# The association between low-grade proteinuria and adverse kidney outcomes in IgA nephropathy: A systematic review and meta-analysis

**DOI:** 10.1101/2025.08.12.25333541

**Authors:** Yuya Yamaguchi, Takaaki Kosugi, Takaya Sasaki, Kotaro Haruhara, Yusuke Okabayashi, Masahiro Okabe, Akihiro Shimizu, Shinya Yokote, Sradha Kotwal, Min Jun, Brendon L Neuen, Kazuhiko Tsuruya, Hiroyuki Ueda, Nobuo Tsuboi, Takashi Yokoo

## Abstract

**Background:** Overt proteinuria (>1.0 g/day) is a well-established risk factor for kidney disease progression in IgA nephropathy (IgAN). However, recent evidence suggests that even low-grade proteinuria, typically defined as 0.5–1.0 g/day, may be clinically significant. The prognostic impact of low-grade proteinuria has not been systematically evaluated.

**Methods:** We conducted a systematic review and meta-analysis to evaluate the association between low-grade proteinuria and adverse kidney outcomes in patients with IgAN. A systematic literature search was performed on PubMed and Web of Science. Eligible studies included those reporting on kidney outcomes such as estimated glomerular filtration rate (eGFR) decline, kidney failure, or eGFR slope in relation to low-grade proteinuria measured either at baseline or during follow-up (e.g., time-averaged proteinuria [TAP]). Data were synthesized using random-effects meta-analysis. No funding was received for this review. The protocol was registered in OSF REGISTRIES [https://osf.io/5dfqr].

**Results:** A total of 23 studies involving 15,289 patients met the inclusion criteria. Baseline low-grade proteinuria was significantly associated with an increased risk of adverse kidney outcomes compared to proteinuria below 0.5 g/day (pooled hazard ratio [HR] 1.73, 95% confidence interval [CI] 1.36–2.20). Similarly, low-grade TAP was associated with a higher risk of kidney outcomes (pooled HR 2.87, 95% CI 1.48–5.56) and a significantly steeper annual decline in eGFR (mean difference −1.02 mL/min/1.73 m^2^/year, 95% CI −1.60 to −0.45). Subgroup analyses based on geographic region and leave-one-out sensitivity analyses were consistent with the overall findings.

**Conclusions:** Low-grade proteinuria, whether assessed at baseline or over time, is an important predictor of kidney disease progression in patients with IgAN. These results reinforce recent clinical guidelines recommending proteinuria control under 0.5 g/day. Long-term suppression of proteinuria should be considered a key therapeutic goal in IgAN.

## Introduction

Immunoglobulin A nephropathy (IgAN) is the most common primary glomerulonephritis worldwide and a leading cause of chronic kidney disease (CKD) and kidney failure, particularly in adolescents and young adults [1]. The clinical course of IgAN is highly variable, ranging from isolated hematuria to progressive kidney function decline. Among established prognostic markers, proteinuria is consistently identified as a strong and independent risk factor for adverse kidney outcomes, including estimated glomerular filtration rate (eGFR) decline and progression to kidney failure [2].

While the clinical importance of overt proteinuria (>1.0 g/day) is well recognized, emerging evidence suggests that even low-grade proteinuria, defined as 0.5–1.0 g/day, is associated with a significantly increased risk of kidney outcomes [3, 4]. In East Asian countries, clinicians have long regarded this lower range of proteinuria as clinically meaningful [5, 6], and Japanese clinical guidelines have historically set 0.5 g/day as the threshold for initiating interventions [7]. More recently, large-scale epidemiologic studies including Western populations have demonstrated similar findings, indicating that low-grade proteinuria is associated with a higher risk of kidney failure [8, 9].

Reflecting this evolving understanding, the 2021 Kidney Disease: Improving Global Outcomes (KDIGO) guidelines recognize proteinuria reduction as a surrogate marker of improved kidney outcomes in IgAN and recommend targeting levels below 1.0 g/day as a reasonable therapeutic goal [4]. The upcoming KDIGO guidelines for IgAN are expected to reinforce this recommendation [10], potentially redefining treatment goals for a large subset of patients. Despite these shifts, current recommendations are primarily based on individual observational studies and narrative syntheses. No prior systematic review and meta-analysis has specifically focused on the risk for kidney outcomes associated with low-grade proteinuria in IgAN.

To address this evidence gap, the present systematic review and meta-analysis aimed to evaluate the prognostic impact of low-grade proteinuria (0.5–1.0 g/day or g/g) in patients with IgAN, compared with minimal proteinuria, to inform clinical decision-making and future guideline development.

## Methods

### Study design and registration

This systematic review and meta-analysis was conducted in accordance with the MOOSE guidelines. The review protocol was developed prior to initiation and registered with OSF REGISTRIES (registration ID: 5dfqr [https://osf.io/5dfqr]).

### Eligibility criteria

We included observational studies and clinical trials reporting kidney outcomes in patients with IgAN and low-grade proteinuria. Studies were eligible if they met the following inclusion criteria: the population comprised patients with IgAN; the exposure was low-grade proteinuria, typically defined as 0.5–1.0 g/day or equivalent units (e.g., urinary protein-to-creatinine ratio [UPCR], g/g). Proteinuria during the follow-up of the study (e.g., time-averaged proteinuria [TAP], time-weighted average proteinuria [TWAP], and time-varying proteinuria [TVP]) was considered eligible as well as baseline proteinuria. The definitions of proteinuria over time were as follows: TAP, the arithmetic mean of proteinuria values measured at multiple time points during follow-up; TWAP, the average proteinuria level calculated by weighting each measurement according to the duration between time points; and TVP, a time-dependent variable representing proteinuria levels at various time points. Studies reporting albuminuria were also eligible, and the cut-off values for proteinuria were converted to those of urinary albumin-to-creatinine ratio (UACR) using established conversion formulas [11]. When the exposure range did not exactly match the target group (0.5–1.0 g/day), we allowed for an approximate 50% deviation, including studies in which the exposure range extended from a minimum of 0.25 g/day to a maximum of 1.5 g/day. The comparator was either minimal or no proteinuria, typically <0.5 g/day, depending on the study design. Very low-grade proteinuria, defined as a category between minimal and low-grade proteinuria, was included as an exploratory exposure when reported in the original studies.

Eligible outcomes of interest included kidney related outcomes (i.e., ≥30–50% decline in eGFR, doubling of serum creatinine, progression to kidney failure, annual eGFR slope, or kidney related mortality). The primary outcome was defined as a composite of these events.

Studies were excluded if they were: (1) case reports, cross-sectional studies, reviews, editorials, or expert opinions; (2) inaccessible in full-text form; (3) not reporting exposure of interest for example studies exclusively focused on high-grade proteinuria (>1.0 g/day) without subgroup analysis for lower proteinuria ranges; or (4) not reporting at least outcomes of interest.

### Information sources

A systematic literature search was conducted in the following databases from inception to the most recent search date July 11^th^, 2025: PubMed, and Web of Science. In addition, reference lists of relevant guidelines (e.g., KDIGO) were manually searched. Grey literature was searched through preprint servers prior to evidence synthesis.

### Search strategy

Search strategies are detailed in **Supplementary Item S1**. In summary, search terms included combinations terms for “IgA nephropathy,” “proteinuria,” and “renal outcomes.” A medical librarian at The Jikei University School of Medicine assisted in developing and validating the strategy. We did not contact study authors for additional information.

### Study selection process

Study selection was conducted using Rayyan (https://rayyan.ai/; Rayyan Systems Inc., Cambridge, MA, USA) for blinded screening. After duplicates were removed, two reviewers (Y.Y. and T.K.) independently screened titles and abstracts, then full texts, against the eligibility criteria. Disagreements were resolved by a third reviewer (T.S.).

### Data extraction process

Data were extracted independently by two reviewers (Y.Y. and T.K.) using a standardized Microsoft Excel template (Microsoft Corp., Redmond, WA, USA). Extracted information included study characteristics (author, year, geographic region, sample size); patient demographics (age, sex, eGFR, proteinuria, comorbidities); proteinuria category; number of outcomes; and effect estimates (hazard ratios [HR], and mean difference [MD] with 95% confidence intervals [CI]). Discrepancies were resolved by consensus or arbitration by a third reviewer (T.S.).

### Quality assessment

Risk of bias was assessed using the Newcastle-Ottawa Scale (NOS), considering selection, comparability, and outcome domains. Two reviewers (Y.Y. and T.K.) independently evaluated NOS. Discrepancies were resolved by a third reviewer (T.S.). Certainty of evidence for each primary outcome was assessed using the GRADE approach, considering risk of bias, inconsistency, indirectness, imprecision, and publication bias.

### Statistical analysis

Where applicable, meta-analyses were performed using a random-effects model. Pooled effect estimates (e.g., HRs) were calculated, and heterogeneity was assessed using the *I^2^* statistic and Cochran’s Q test. If fewer than three studies were available or data were not suitable for quantitative synthesis, a narrative synthesis was conducted. Subgroup analyses by geographic region (Asian vs. non-Asian) were also performed to examine potential effect modification. For sensitivity analyses, leave-one-out analyses were performed to test robustness. Publication bias was planned to be assessed using funnel plots and Egger’s test when at least 10 studies were available; however, this was not conducted due to an insufficient number of studies. All statistical analyses were performed using R version 4.5.0 (R Foundation, Vienna, Austria).

## Results

### Study selection and characteristics

As shown in **Figure 1**, a total of 1,891 articles were identified. After removing duplicates, 1,419 abstracts were screened. Of these, 183 articles were selected for full-text review, and ultimately 23 studies met the inclusion criteria and were included in the present review [6, 8, 9, 12–31]. No eligible studies were excluded despite meeting the inclusion criteria.

**Figure 1.**
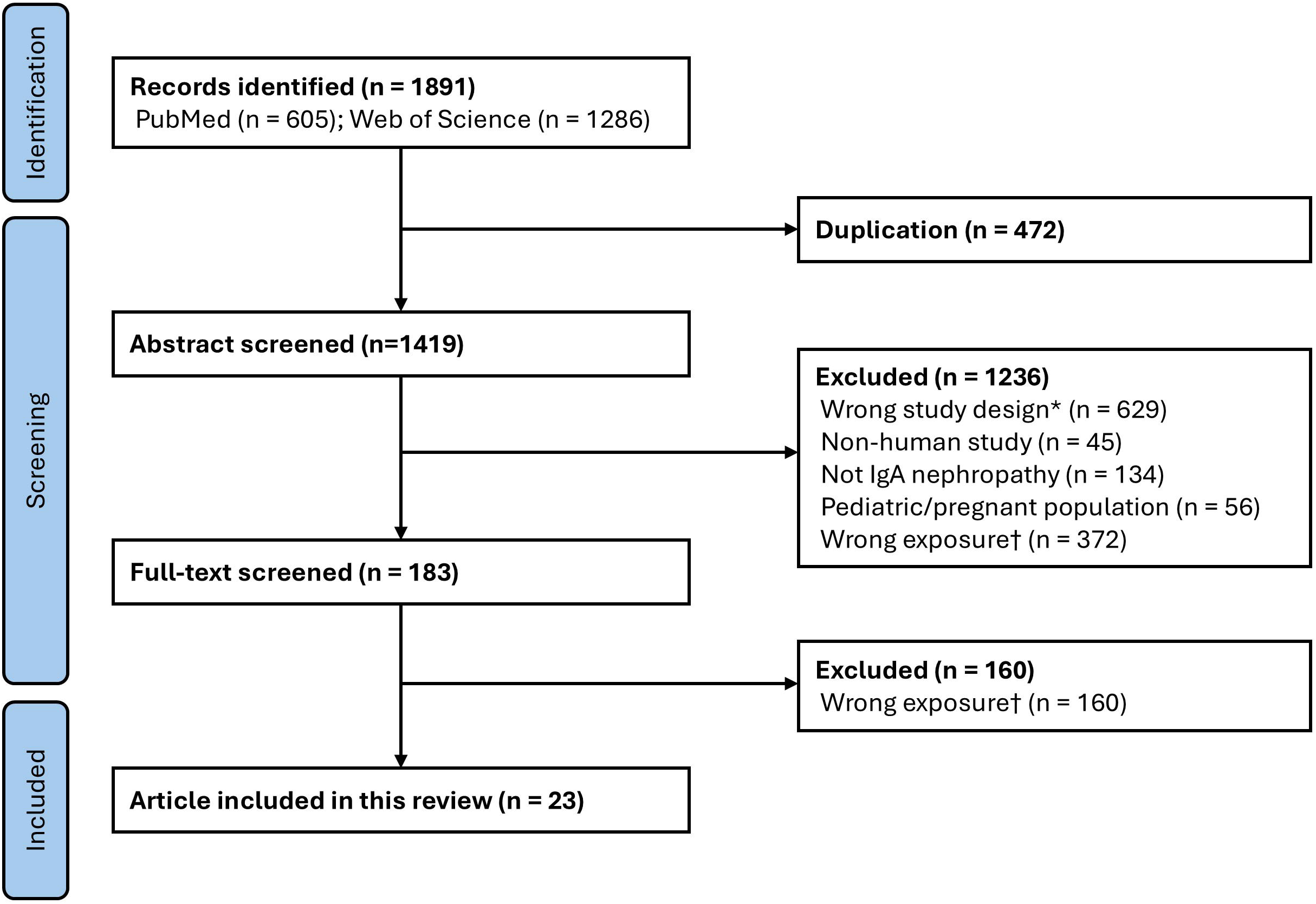
Flow diagram of study selection process. Flowchart outlining the identification, screening, eligibility, and inclusion of studies in this systematic review. A total of 23 articles were ultimately included. References in the KDIGO guidelines [4, 10] were hand-searched, but no additional studies were added. One hundred and thirty-eight pre-print articles from medRxiv were identified and underwent full-text screening prior to integration, but all were excluded.

The characteristics of the included studies are summarized in **Table 1** and **Supplementary Table S1**. These studies were conducted across diverse geographical regions, including Asia (China, India, Japan, Korea, Malaysia), Europe (Italy, Switzerland, Sweden, Germany, United Kingdom), and North America (United States, Canada).

**Table 1.**
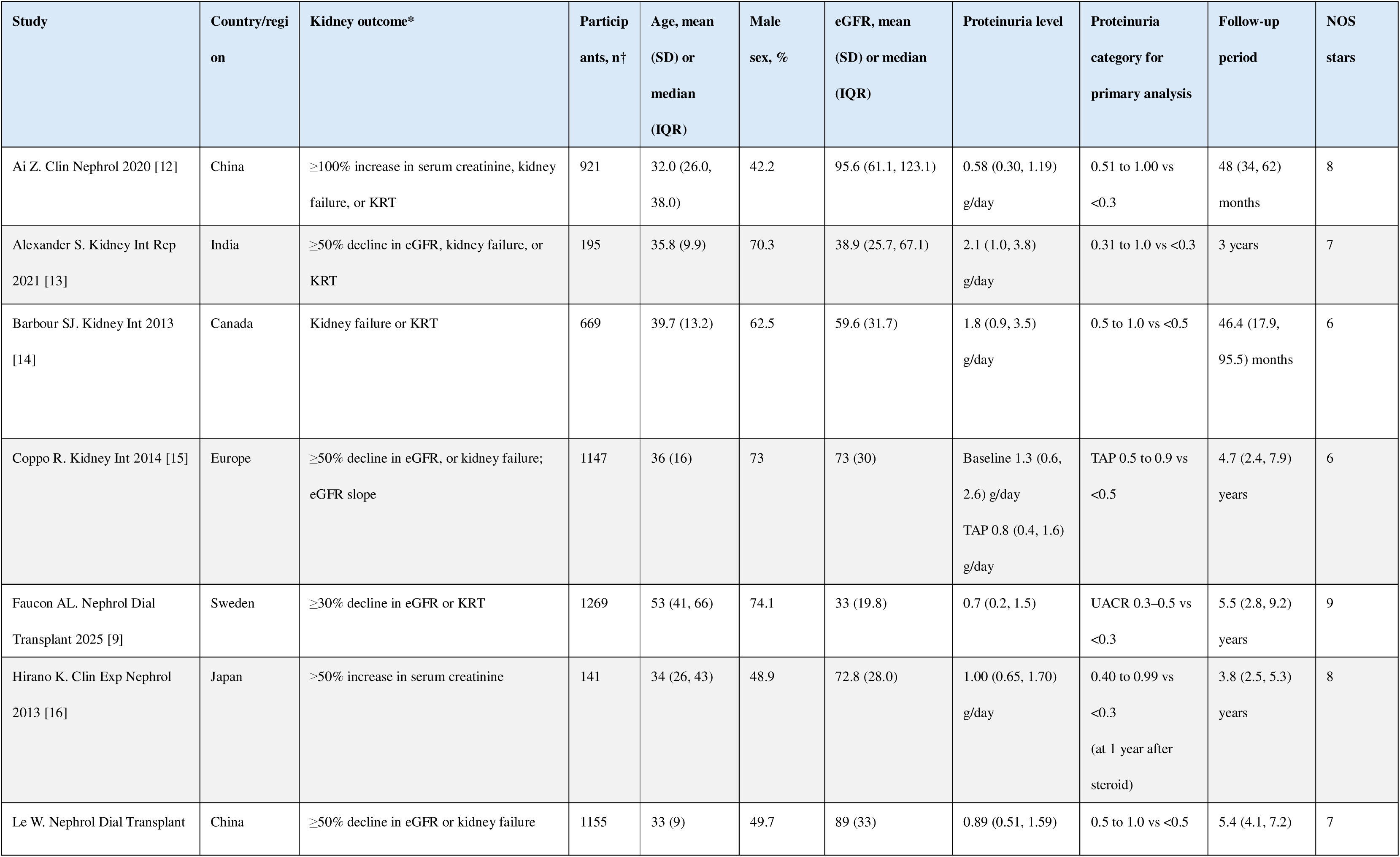
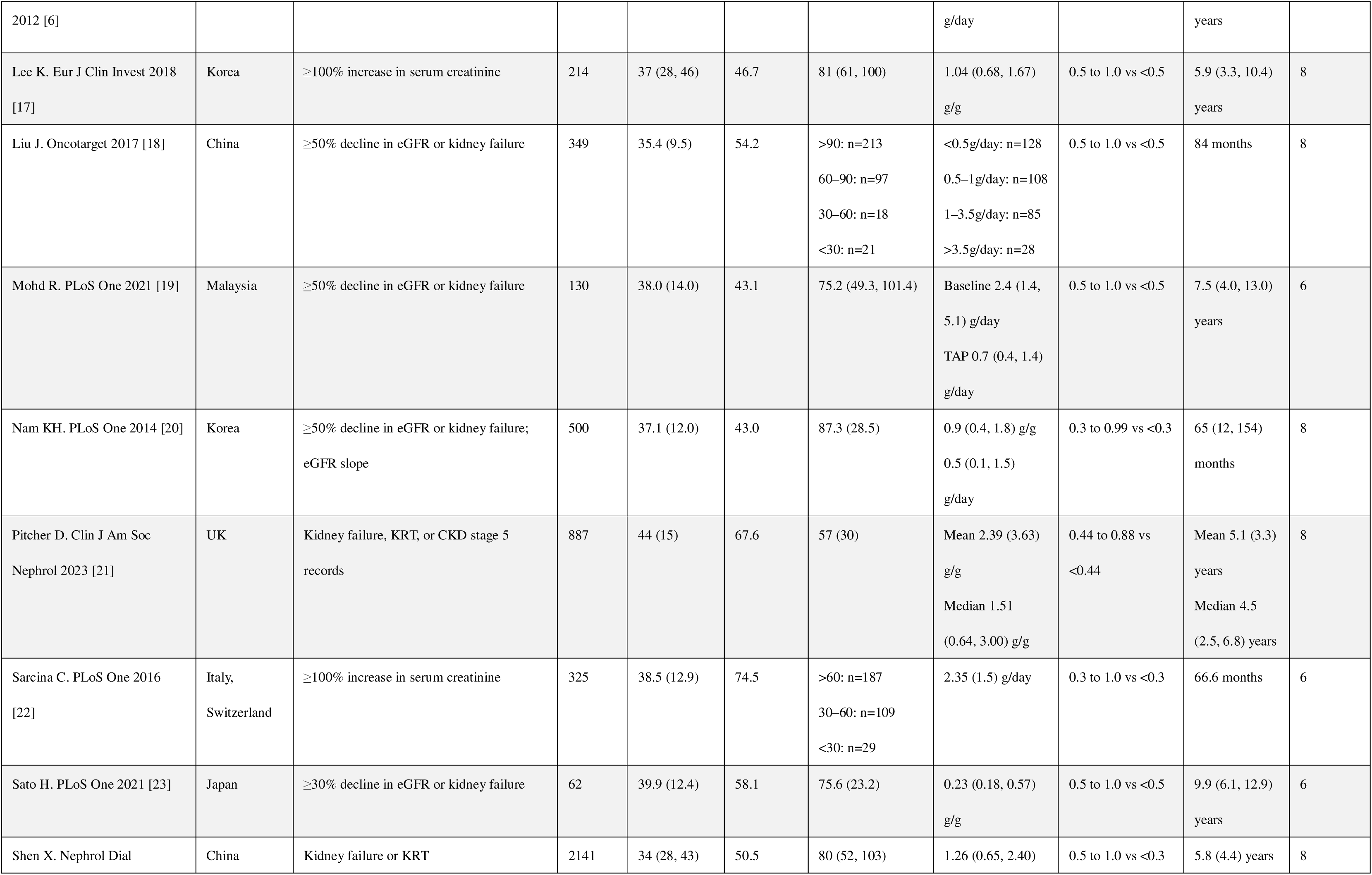
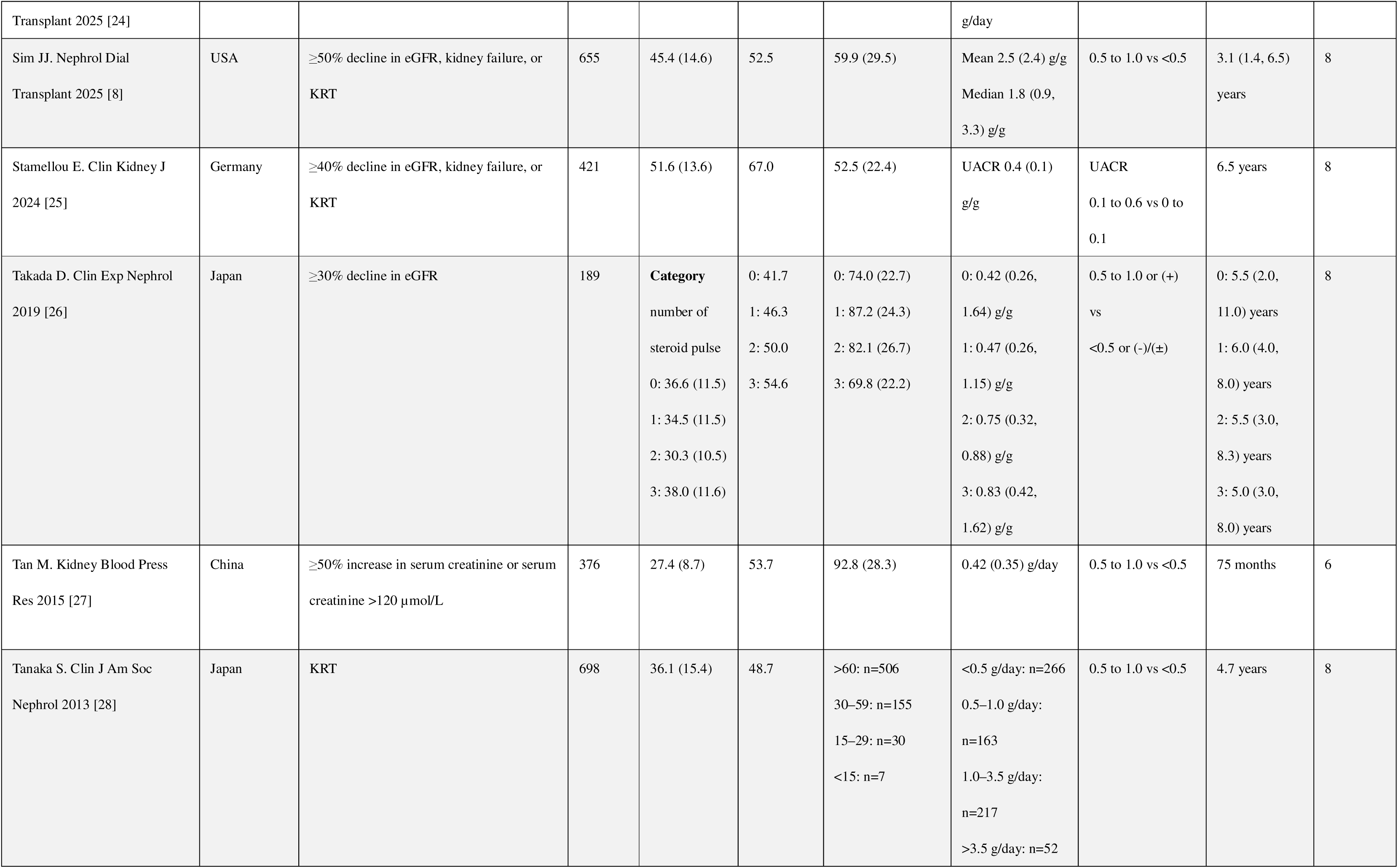
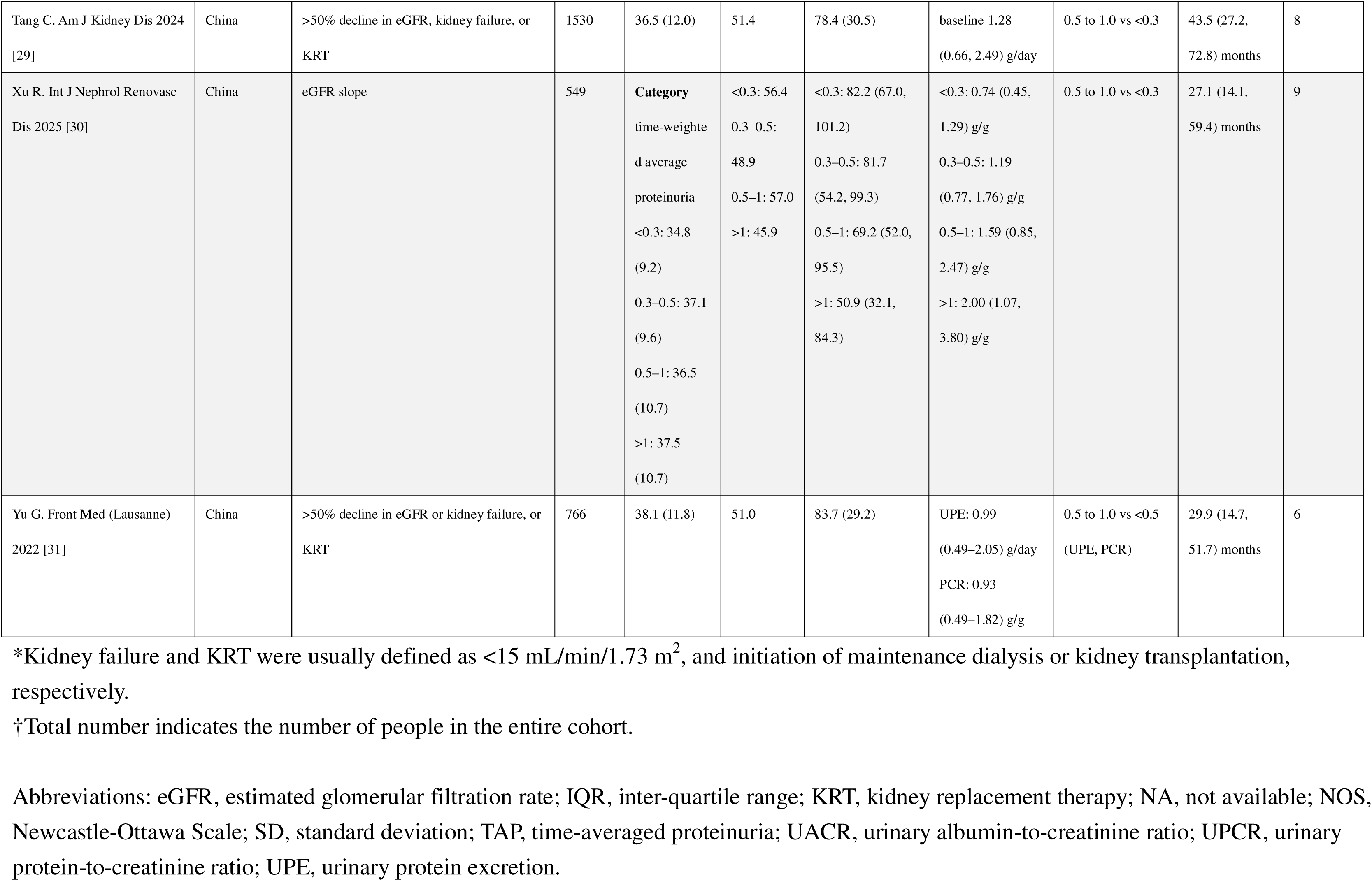
Study characteristics included in the present systematic review (n = 23).

Sample sizes ranged from 62 to 2,141 patients, with a cumulative total of 15,289 patients. The mean or median age of participants varied from 27 to 54 years across studies, and the proportion of male participants ranged from 42% to 75%.

Baseline kidney function was reported using eGFR, which ranged from approximately 33 to 96 mL/min/1.73 m^2^. Proteinuria was assessed at baseline using either 24-hour urine protein excretion, UPCR, or UACR. Studies also examined TAP, TWAP or TVP as exposure variables.

The adverse kidney outcomes considered in the present review included doubling of serum creatinine, ≥30–50% decline in eGFR, kidney failure, kidney replacement therapy, and eGFR slope. Follow-up periods ranged from 3.1 to over 9 years.

Regarding treatment, the proportion of patients receiving glucocorticoids or other immunosuppressive agents varied substantially between studies depending on the study population and design. The use of renin–angiotensin system inhibitors was reported in the most studies, with usage rates ranging from 53% to 96% among the studies that provided such data. Use of sodium-glucose co-transporter-2 inhibitors was infrequently reported, as most studies predate their widespread adoption.

Risk of bias was assessed using the NOS. Sixteen studies were rated as good quality (NOS stars: ≥7), while the remaining seven were rated as fair quality (NOS stars: 4 to 6). Detailed assessments of NOS are provided in **Supplementary Table S2**.

### Baseline proteinuria

A total of 15 studies [8, 9, 12–16, 19, 22, 23, 25–28, 31] investigated the association between low-grade baseline proteinuria and adverse kidney outcomes. Among these, 6 studies [13, 15, 19, 23, 27, 31] reported only Kaplan–Meier curves or relative risks and were excluded from the quantitative synthesis. The remaining 9 studies [8, 9, 12, 14, 16, 22, 25, 26, 28] reported HRs comparing low-grade baseline proteinuria with the lowest baseline proteinuria category, with HRs ranging from 0.80 to 4.00. Incorporating these studies into the meta-analysis, pooled analysis showed that low-grade baseline proteinuria was significantly associated with an increased risk of adverse kidney outcomes (pooled HR 1.73 [95% CI 1.36 to 2.20]; **Figure 2**). No statistically significant heterogeneity was detected.

**Figure 2.**
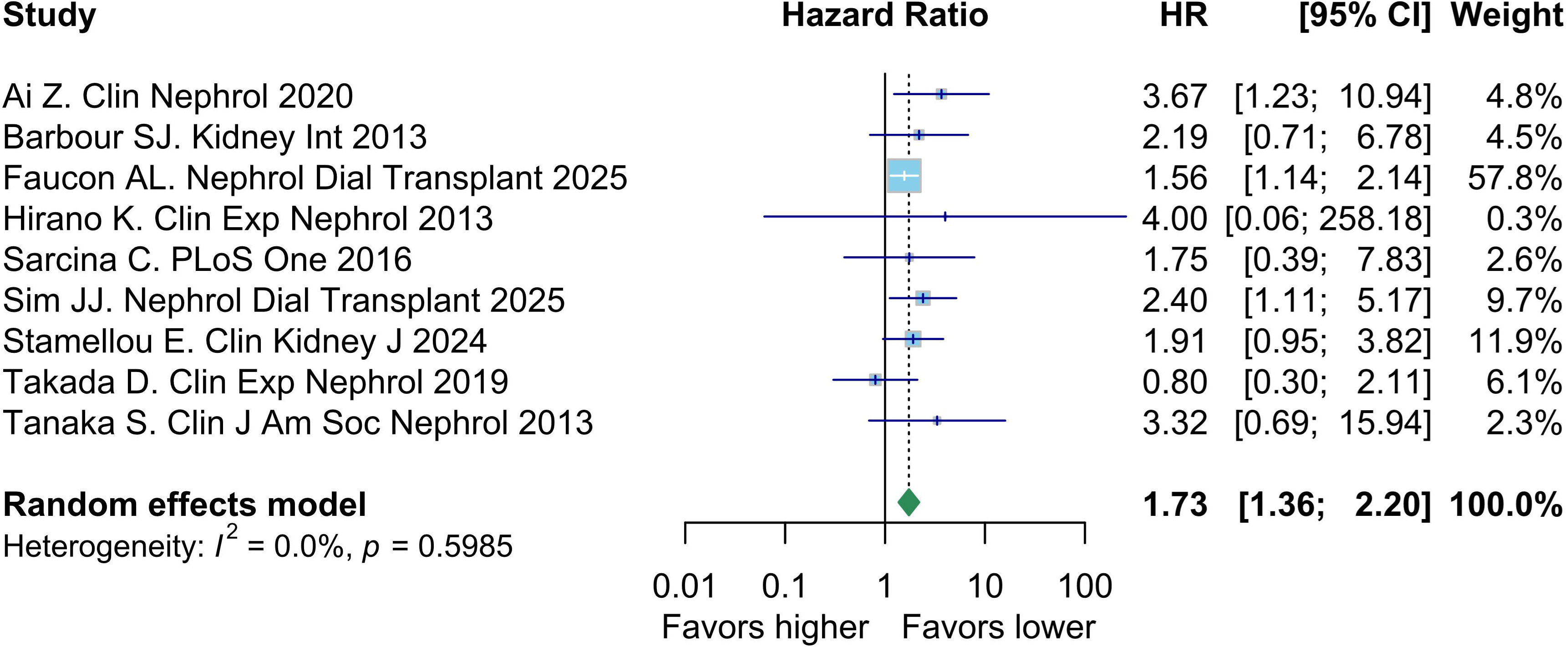
Association between baseline proteinuria and adverse kidney outcomes. Forest plot shows the hazard ratios (HRs) and 95% confidence intervals (CIs) for the risk of baseline proteinuria for adverse kidney outcomes across 9 cohort studies. The meta-analysis was performed using a random-effects model.

Regarding eGFR slope, only one study reported this outcome, but its standard deviations were not described [25].

### Time-averaged proteinuria

Among the included studies, 8 studies assessed the risk of TAP for kidney outcomes. Of these, 7 studies [6, 8, 17, 18, 20, 21, 24] reported HRs for the association between low-grade TAP and adverse kidney outcomes, with HRs ranging from 1.07 to 9.10. The pooled analysis demonstrated that low-grade TAP was associated with a significantly increased risk of adverse kidney outcomes (pooled HR 2.87 [95% CI 1.48 to 5.56]; **Figure 3**). Substantial heterogeneity was observed across studies (*I^2^*= 77.1%).

**Figure 3.**
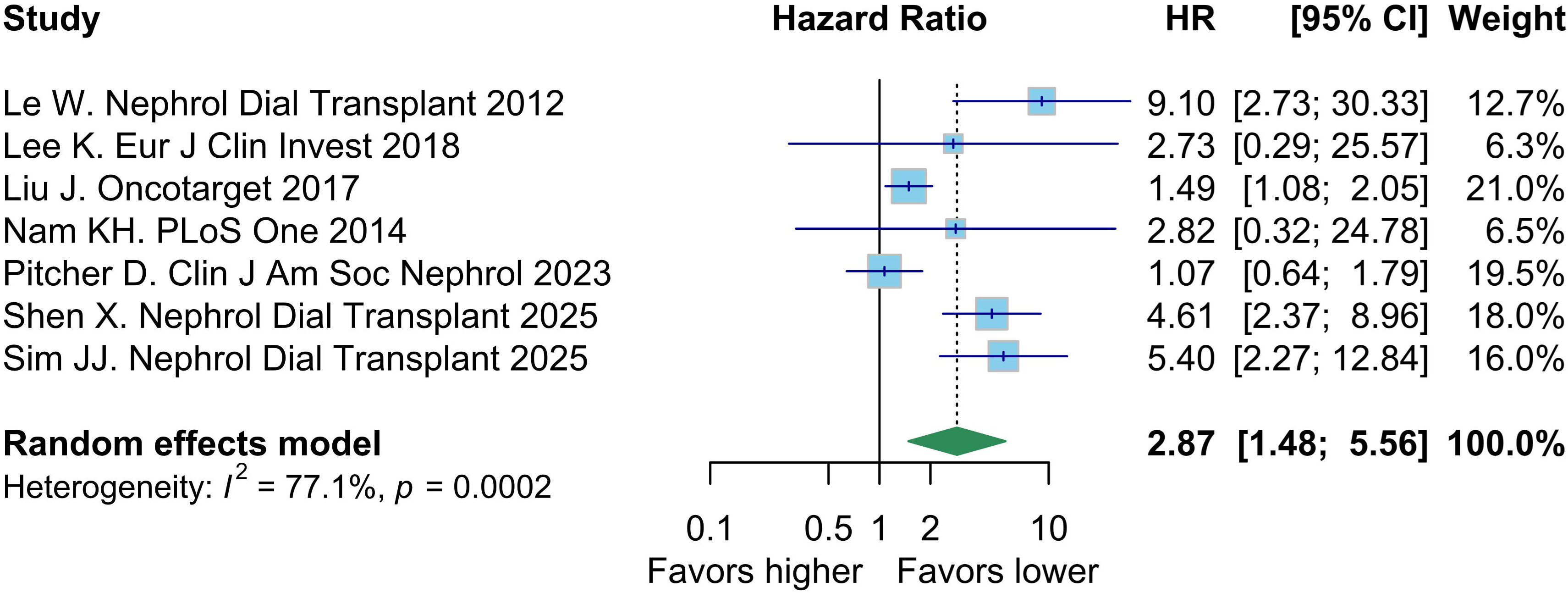
Association between time-averaged proteinuria (TAP) and adverse kidney outcomes. Forest plot shows the hazard ratios (HRs) and 95% confidence intervals (CIs) for the risk of TAP for adverse kidney outcomes across 7 cohort studies. The meta-analysis was performed using a random-effects model.

For eGFR slope, 5 studies [6, 20–22, 24] provided relevant data. After excluding one study [22] due to missing standard deviation, the meta-analysis showed that low-grade TAP was associated with a significantly faster eGFR decline compared to the lowest TAP categories (pooled mean difference −1.02 [95% CI −1.60 to −0.45] mL/min/1.73 m^2^/year; **Figure 4**) [6, 20, 21, 24]. Substantial heterogeneity was observed across studies (*I^2^* = 80.1%).

**Figure 4.**
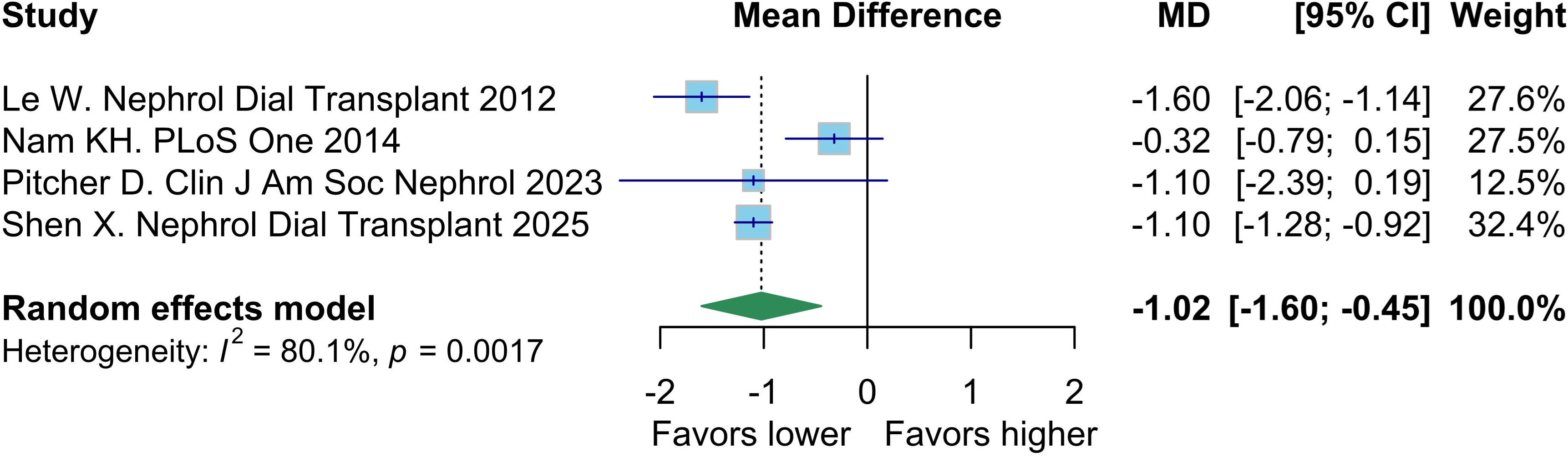
Association between time-averaged proteinuria (TAP) and annual estimated glomerular filtration rate slope. Forest plot shows the mean differences (MDs) and 95% confidence intervals (CIs) for the influence of TAP on eGFR slope outcomes across 4 cohort studies. The meta-analysis was performed using a random-effects model.

### Time-weighted average proteinuria

Only one study [30] reported on TWAP and eGFR slope, with no analysis evaluating the influence of TWAP on adverse kidney outcomes. In patients with baseline eGFR 15–60 mL/min/1.73 m^2^, low-grade TWAP was associated with a steeper eGFR decline (−3.38 [95% CI −5.12 to −1.65] mL/min/1.73 m^2^/year). However, no significant association was found among patients with eGFR >60 mL/min/1.73 m^2^ (0.52 [95% CI −0.48 to 1.52] mL/min/1.73 m^2^/year).

### Time-varying proteinuria

One study [29] assessed the influence of TVP on adverse kidney outcomes. The results revealed that low-grade TVP was significantly associated with increased risk of adverse kidney outcomes compared to no or minimal TVP (HR 4.04 [95% CI 1.93 to 8.46]).

### Subgroup analysis and sensitivity analysis

Subgroup analyses by geographic region (Asian vs. non-Asian) showed no significant heterogeneity between subgroups across any of the comparisons (**Supplementary Figures S1–S3**). To assess the robustness of the findings, leave-one-out sensitivity analyses were performed for each meta-analysis (**Supplementary Tables S3–S5**), revealing that no single study had a disproportionate impact on the overall effect estimates.

### Very low-grade proteinuria

Five studies [12, 16, 24, 29, 30] reported outcomes for subcategories of very low-grade proteinuria, typically between the low-grade category and the lowest category (**Table 2**). These results were heterogeneous in definition and reporting format. Ai, *et al.* reported significantly increased HR of very low-grade baseline proteinuria [12]. Xu, *et al.* revealed a significantly steeper annual eGFR slope of very low-grade TWAP among the decreased kidney function [30]. Remaining studies reported non-significant results.

**Table 2.** Findings from subcategories of very low-grade proteinuria.

| ***(A) Adverse kidney outcomes*** |  |  |  |  |  |
| --- | --- | --- | --- | --- | --- |
| **Study** | **Proteinuria index** | **Definition of very low-grade proteinuria** | **Definition of reference** | **Participants, n** | **HR (95%CI)** |
| Ai Z, et al. Clin Nephrol 2020 | Baseline proteinuria | 0.3–0.5 g/day | <0.3 g/day | 158 | 3.70 (1.09, 12.56) |
| Hirano K, et al. Clin Exp Nephrol 2013 | Baseline proteinuria | 0.3–0.4 g/day | <0.3 g/day | 23 | 0.33 (0.01, 11.10) |
| Shen X, et al. Nephrol Dial Transplant 2025 | TAP | 0.3–0.5 g/day | <0.3 g/day | 296 | 1.05 (0.43, 2.70) |
| Tang C, et al. Am J Kidney Dis 2024 | TVP | 0.3–0.5 g/day | <0.3 g/day | NA | 2.22 (0.88, 5.58) |
| ***(B) Annual eGFR slope, mL/min/1.73 m<sup>2</sup>/year*** |  |  |  |  |  |
| **Study** | **Proteinuria index** | **Definition of very low-grade proteinuria** | **Definition of reference** | **Participants, n** | **MD (95%CI)** |
| Xu R et al. Int J Nephrol Renovasc Dis 2025 (eGFR 15–60 mL/min/1.73 m<sup>2</sup>) | TWAP | 0.3–0.5 g/g | <0.3 g/g | NA | -2.04 (-3.72, -0.35) |
| Xu R, et al. Int J Nephrol Renovasc Dis 2025 (eGFR >60 mL/min/1.73 m<sup>2</sup>) | TWAP | 0.3–0.5 g/g | <0.3 g/g | NA | -0.04 (-1.98, 1.89) |

Abbreviations: eGFR, estimated glomerular filtration rate; HR, hazard ratio; MD, mean difference; TAP, time-averaged proteinuria; TVP, time-varying proteinuria; TWAP, time-weighted average proteinuria

### Certainty of evidence

The certainty of evidence was assessed using the GRADE approach and is summarized in **Table 3**. The certainty of the evidence regarding baseline proteinuria and adverse kidney outcomes was rated as moderate, with a low risk of bias, no serious inconsistency, and consistent effects observed across subgroup and sensitivity analyses. Similarly, the evidence for TAP’s association with adverse kidney outcomes and with annual eGFR slope was also rated as moderate certainty. This was supported by low risk of bias, consistent findings despite moderate heterogeneity, and indications of a large or possibly large effect size.

**Table 3.** Certainty of each evidence based on GRADE.

| Exposure | Outcome | Downgrade domain |  |  |  |  | Upgrade domain |  |  | Certainty | Comments for judgement |
| --- | --- | --- | --- | --- | --- | --- | --- | --- | --- | --- | --- |
|  |  | Risk of bias (RoB) | Inconsistency | Indirectness | Imprecision | Publication bias | Large effect | Dose response gradient | Residual confounding minimizing the effect |  |  |
| Baseline proteinuria | Adverse kidney outcomes | Not serious | Not serious | Not serious | Not serious | Not applicable (no downgrade) | No | Yes | No | ⊠⊠⊠⊠<br>moderate | - Low RoB<br>- No heterogeneity<br>- Consistent effect across leave-one-out sensitivity analyses<br>- Established higher proteinuria effect |
| Time-averaged proteinuria | Adverse kidney outcomes | Not serious | Not serious | Not serious | Not serious | Not applicable (no downgrade) | Yes | Yes | No | ⊠⊠⊠⊠<br>moderate | - Low RoB<br>- Moderate heterogeneity but consistent effect across leave-one-out sensitivity analyses<br>- Large effect size<br>- Established higher proteinuria effect |
| Time-averaged proteinuria | Annual eGFR slope | Not serious | Not serious | Not serious | Not serious | Not applicable (no downgrade) | Possibly | Yes | No | ⊠⊠⊠⊠<br>moderate | - Low RoB<br>- Moderate heterogeneity but consistent effect across leave-one-out sensitivity analyses<br>- Possibly large effect size<br>- Established higher proteinuria effect |

Abbreviation: eGFR, estimated glomerular filtration rate.

## Discussion

This systematic review and meta-analysis demonstrated that low-grade proteinuria, typically defined as 0.5–1.0 g/day or g/g, in patients with IgAN is associated with a significantly increased risk of adverse kidney outcomes and steeper annual eGFR slope. This association was generally consistent across different measures of proteinuria (i.e., baseline proteinuria, TAP, TWAP and TVP), and across diverse study characteristics, and remained robust across subgroup and sensitivity analyses.

Our analysis shows that both baseline proteinuria and longitudinal proteinuria measures (i.e., TAP, TWAP, and TVP) are independently associated with adverse kidney outcomes and annual eGFR slope in IgAN [6, 8, 9, 12–31]. Baseline proteinuria is considered valuable at the time of diagnosis for initial risk assessment, guiding early prognostic estimation and assessment of the need for intervention. Furthermore, the strong association of TAP and TVP with adverse kidney outcomes suggests that the cumulative exposure to proteinuria over time may be more relevant than baseline proteinuria. These findings shed light on the importance of achieving a reduction in proteinuria and sustaining suppression of proteinuria throughout the disease course. In this context, therapeutic interventions aimed at maintaining lower proteinuria levels long-term are critical for modifying disease trajectory. This supports a treat-to-target strategy, where therapy is continued or adjusted to maintain proteinuria below 0.5 g/day.

Our findings underscore that low-grade proteinuria (0.5–1.0 g/day) should not be considered benign in patients with IgAN. These patients may benefit from closer monitoring and should be considered appropriate candidates for therapeutic intervention. Importantly, recent clinical trials in IgAN have largely focused on patients with proteinuria >1.0 g/day [32–36], thereby excluding a substantial subset of patients with potentially meaningful risk. Future trials should consider including patients with low-grade proteinuria to evaluate treatment efficacy and benefit in this underrepresented group.

Few studies have reported on very low-grade proteinuria (e.g., 0.3–0.5 g/day) [12, 16, 24, 29, 30], with inconsistent findings. While some suggested an increased risk of kidney outcomes even at this level, others found no significant association—possibly due to limited statistical power or varying definitions. These inconsistencies leave the prognostic significance of very low-grade proteinuria uncertain, highlighting the need for more targeted and adequately powered research.

Our findings are consistent with and reinforce the 2021 KDIGO recommendation to target proteinuria levels below 1.0 g/day in patients with IgAN [4]. The results provide quantitative support for lowering treatment thresholds and may inform future KDIGO updates [10], particularly regarding risk stratification and intervention timing in patients with low-grade proteinuria.

This study has several limitations. All included studies were observational, limiting causal inference. Variations in the definitions of proteinuria and kidney outcomes may have introduced potential misclassification. Substantial heterogeneity was observed in the TAP-based meta-analyses (*I^2^* >75%). This heterogeneity likely reflects differences in treatment strategies, follow-up durations, and how TAP was defined across studies. Nonetheless, the direction of association remained consistent, and we used a random-effects model to account for between-study variability. Leave-one-out sensitivity analyses confirmed the robustness of results, indicating that no single study disproportionately influenced the overall estimates. Additionally, subgroup analyses based on geographic region (Asian vs. non-Asian) showed no significant differences, supporting the consistency of findings across populations. Finally, publication bias could not be formally evaluated due to the limited number of eligible studies for some outcomes; however, the consistency of results across sensitivity and subgroup analyses suggests that the impact of potential publication bias may be limited.

In conclusion, this systematic review and meta-analysis show that low-grade proteinuria in IgAN is associated with a significantly increased risk of kidney disease progression. These findings challenge the notion that proteinuria below 1.0 g/day is low risk and suggest that closer monitoring and possibly earlier treatment may be warranted in patients with low-grade proteinuria. Future interventional studies should include this patient population to guide evidence-based care across the full spectrum of IgAN severity.

## Data Sharing Statement

This is a systematic review and meta-analysis; all data used in the analyses are extracted from previously published studies and are fully presented in the manuscript and supplementary materials.

## Acknowledgements

The authors would like to thank the medical librarian in The Jikei University School of Medicine for creating and validating our search strategies.

## Author contribution

Conceptualization and study design: T.S.

Literature search and screening: Y.Y., T.K., T.S., and K.H.

Data extraction: Y.Y., T.K., and T.S.

Risk of bias assessment: Y.Y., T.K., and T.S.

Data analysis and statistical interpretation: Y.Y., T.K., and T.S.

Drafting of the manuscript: T.S.

Critical revision of the manuscript: All authors.

Supervision and project administration: T.S., H.U. N.T., and T.T.

Y.Y. and T.K. contributed equally to this work.

## Supplementary material

Supplementary Item S1. Search strategies for PubMed (A) and Web of Science (B).

Supplementary Figure S1. Subgroup analysis by geographic region (Asian vs. non-Asian) examining the association between baseline proteinuria and adverse kidney outcomes.

Supplementary Figure S2. Subgroup analysis by geographic region (Asian vs. non-Asian) examining the association between time-averaged proteinuria and adverse kidney outcomes.

Supplementary Figure S3. Subgroup analysis by geographic region (Asian vs. non-Asian) examining the association between time-averaged proteinuria and annual estimated glomerular filtration rate slope.

Supplementary Table S1. Supplementary study characteristics.

Supplementary Table S2. Detailed scores of Newcastle-Ottawa Scale (NOS).

Supplementary Table S3. Leave-one-out analyses for association between baseline proteinuria and adverse kidney outcomes.

Supplementary Table S4. Leave-one-out analyses for association between time-averaged proteinuria and adverse kidney outcomes.

Supplementary Table S5. Leave-one-out analyses for association between time-averaged proteinuria and annual estimated glomerular filtration rate slope.

