## Supplemental materials for "The association between low-grade proteinuria and adverse kidney outcomes in IgA nephropathy: A systematic review and meta-analysis"

**<u>Table of contents</u>**

**Supplementary Item S1. Search strategies for PubMed (A) and Web of Science (B).**
**Supplementary Figure S1. Subgroup analysis by geographic region (Asian vs. non-Asian) examining the association between baseline proteinuria and adverse kidney outcomes.**
**Supplementary Figure S2. Subgroup analysis by geographic region (Asian vs. non-Asian) examining the association between time-averaged proteinuria and adverse kidney outcomes.**
**Supplementary Figure S3. Subgroup analysis by geographic region (Asian vs. non-Asian) examining the association between time-averaged proteinuria and annual estimated glomerular filtration rate slope.**
**Supplementary Table S1. Supplementary study characteristics.**
**Supplementary Table S2. Detailed scores of Newcastle-Ottawa Scale (NOS).**
**Supplementary Table S3. Leave-one-out analyses for association between baseline proteinuria and adverse kidney outcomes.**
**Supplementary Table S4. Leave-one-out analyses for association between time-averaged proteinuria and adverse kidney outcomes.**
**Supplementary Table S5. Leave-one-out analyses for association between time-averaged proteinuria and annual estimated glomerular filtration rate slope.**

**Supplementary Item S1. Search strategies for PubMed (A) and Web of Science (B).**

(A) PubMed

| No. | Query | Results |
|---|---|---|
| 1 | ("Glomerulonephritis, IGA"[MeSH Terms] OR "IgA nephropathy"[tiab] OR "Berger* disease"[tiab]) | 10,846 |
| 2 | ( "Proteinuria"[MeSH Terms] OR protein*[tiab] OR "urinary protein"[tiab] OR "urine protein"[tiab] OR "protein excretion"[tiab] OR "UPCR"[tiab] OR "UACR"[tiab] ) | 3,872,544 |
| 3 | ( "kidney outcome*"[tiab] OR "renal outcome*"[tiab] OR "kidney disease progression"[tiab] OR "renal disease progression"[tiab] OR "end-stage renal disease"[tiab] OR "ESRD" [tiab] OR "renal survival"[tiab] OR "kidney survival"[tiab] OR "kidney failure*"[tiab] OR "renal failure*"[tiab] OR "renal prognosis"[tiab] OR "kidney prognosis"[tiab]) | 162,156 |
| 4 | #1 AND #2 AND #3 | 1,356 |
| 5 | (animal[tiab] OR mice[tiab] OR mouse[tiab] OR rat[tiab] OR rats[tiab] OR murin[tiab]) | 3,469,789 |
| 6 | #4 NOT #5 | 1,304 |
| 7 | #4 NOT #6 | 52 |
| 8 | #7 AND "human"[TIAB] | 20 |
| 9 | #6 OR #8 | 1,324 |
| 10 | ("case reports"[pt] OR "review"[pt] OR "meta-analysis"[pt] OR "systematic review"[pt] OR "editorial"[pt] OR "comment"[pt] OR "guideline"[pt] OR "practice guideline"[pt]) | 7,593,856 |
| 11 | #9 NOT #10 | 1,011 |
| 12 | ("Anti-Glomerular Basement Membrane Disease"[MeSH Terms] OR "Glomerulonephritis, Membranoproliferative"[MeSH Terms] OR "Glomerulonephritis, Membranous"[MeSH Terms] OR "Lupus Nephritis"[MeSH Terms] OR "IgA Vasculitis"[MeSH Terms] OR "Diabetic Nephropathies"[MeSH Terms]) | 54,109 |
| 13 | #11 NOT #12 | 917 |
| 14 | ("2010/01/01"[Date - Publication] : "3000"[Date - Publication]) | 18,838,408 |
| 15 | #13 AND #14 | 606 |

(B) Web of Science

| No. | Query | Results |
|---|---|---|
| 1 | TS=("Glomerulonephritis, IGA" OR "IgA nephropathy" OR "Berger* disease") | 10,926 |
| 2 | TS=(Proteinuria OR protein* OR "urinary protein" OR "urine protein" OR "protein excretion" OR UPCR OR UACR) | 4,644,063 |
| 3 | TS=(prognosis OR prognostic OR "disease progression" OR progression OR outcome* OR outcomes OR "end-stage renal disease*" OR ESRD OR "renal survival" OR "kidney survival" OR "kidney failure*" OR "renal failure*") | 4,280,887 |

| | | |
|---|---|---|
| 4 | #3 AND #2 AND #1 | 2,682 |
| 5 | DOP=(2010/2025) | 32,559,236 |
| 6 | #5 AND #4 | 1,830 |
| 7 | TS=(animal OR mice OR mouse OR rat OR rats OR murin) | 3,987,686 |
| 8 | #6 NOT #7 | 1,660 |
| 9 | TS=(human) | 3,989,824 |
| 10 | #6 NOT #8 | 170 |
| 11 | #10 AND #9 | 67 |
| 12 | #11 OR #8 | 1,727 |
| 13 | #11 OR #8 and Article (Document Types) | 1,434 |
| 14 | TS=("Anti-Glomerular Basement Membrane Disease" OR "Glomerulonephritis, Membranoproliferative" OR "Glomerulonephritis, Membranous" OR "Lupus Nephritis" OR "IgA Vasculitis" OR "Diabetic Nephropath*") | 44,866 |
| 15 | #13 NOT #14 | 1,286 |

**Supplementary Figure S1. Subgroup analysis by geographic region (Asian vs. non-Asian) examining the association between baseline proteinuria and adverse kidney outcomes.**

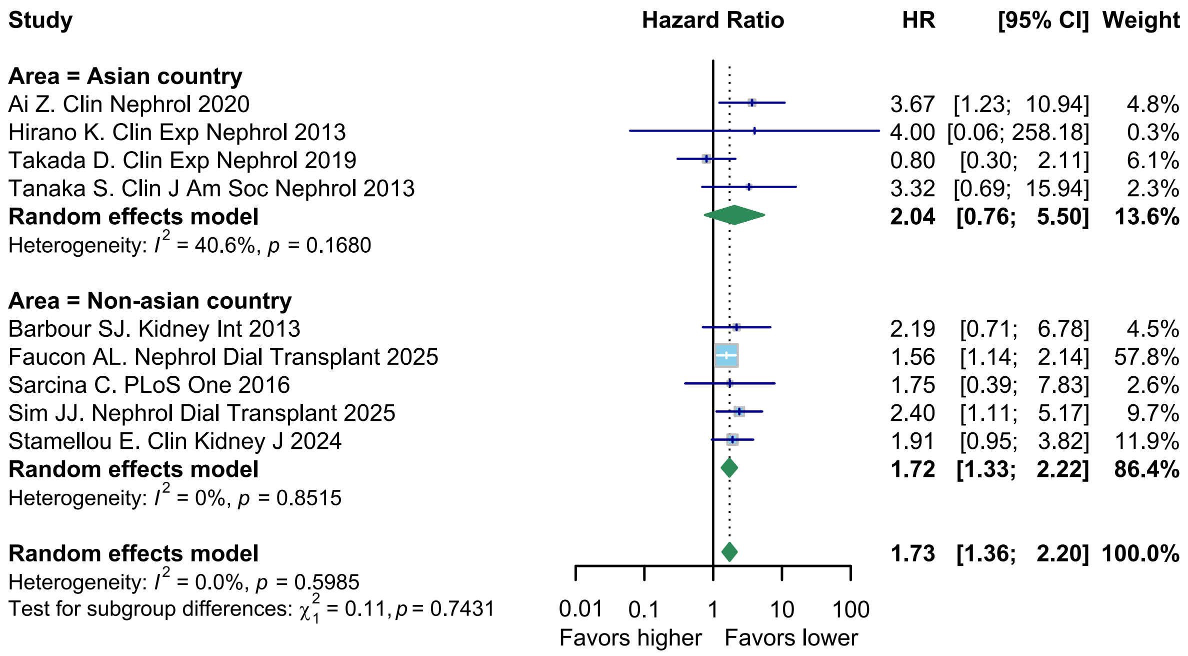

**Supplementary Figure S2. Subgroup analysis by geographic region (Asian vs. non-Asian) examining the association between time-averaged proteinuria and adverse kidney outcomes.**

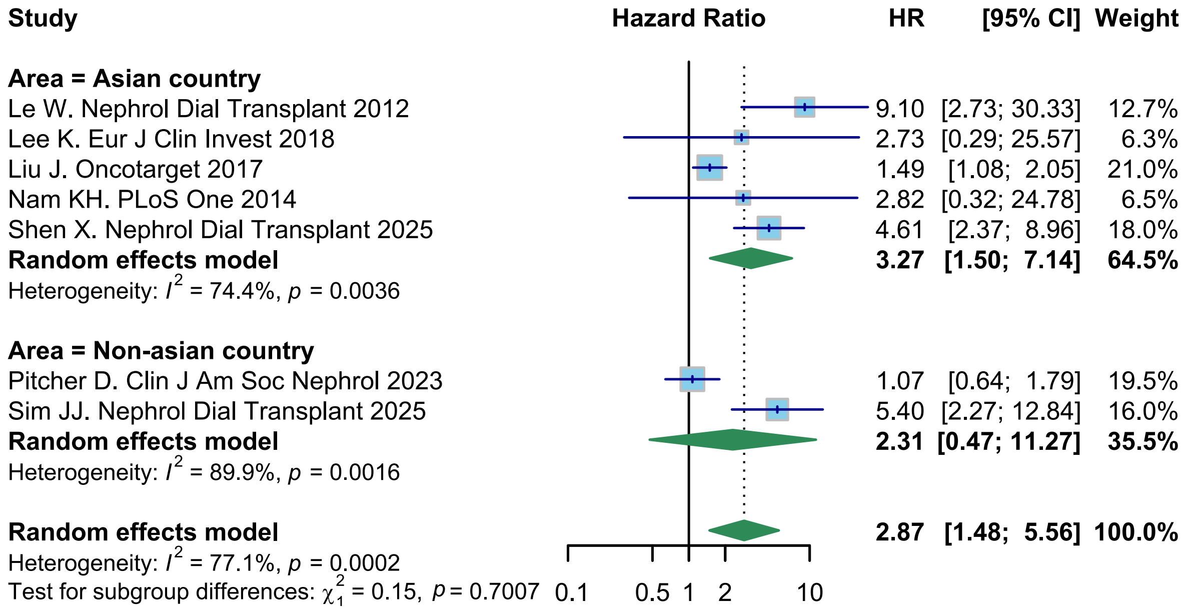

**Supplementary Figure S3. Subgroup analysis by geographic region (Asian vs. non-Asian) examining the association between time-averaged proteinuria and annual estimated glomerular filtration rate slope.**

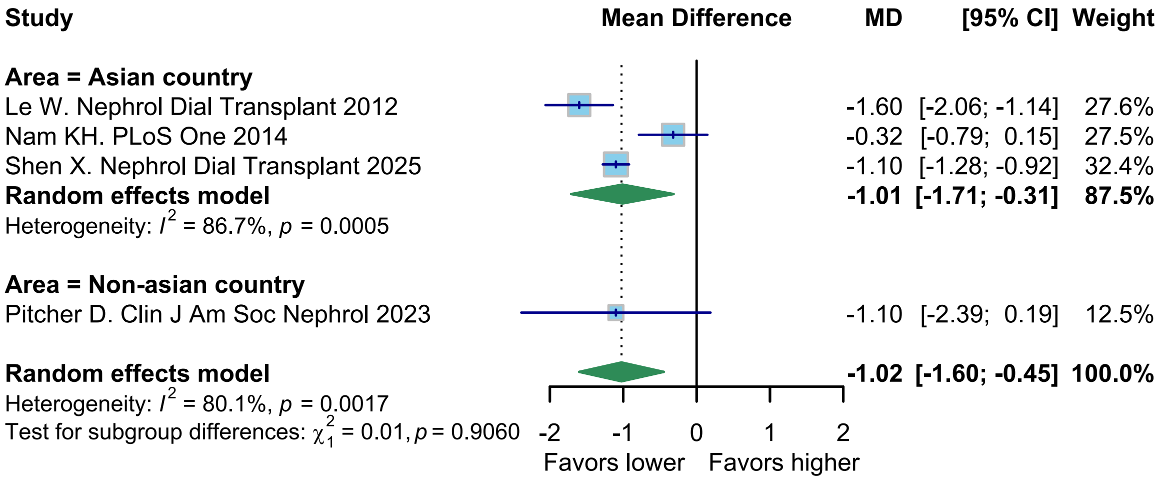

**Supplementary Table S1. Supplementary study characteristics.**

| Study name | Inclusion criteria | Timing of inclusion | Number of participants | | | Baseline and initial treatment during follow-up | | | | |
|---|---|---|---|---|---|---|---|---|---|---|
| | | | Reference (lowest) | Low-grade | Very low-grade | GC, % | IS, % | GC or IS, % | RAS inhibitor, % | SGLT2 inhibitor, % |
| Ai Z. Clin Nephrol 2020 | Patients with biopsy-proven primary IgAN, aged ≥14 years, with eGFR >15 mL/min/1.73 $m^2$ at the time of biopsy, ≥10 glomeruli in the biopsy sample, available 24-hour urine protein data at biopsy, and a follow-up duration ≥12 months | The time of biopsy | 239 | 238 | 158 | 30.4 | NA | NA | 80.4 | NA |
| Alexander S. Kidney Int Rep 2021 | Patients aged ≥18 years with biopsy-proven primary IgA nephropathy who had not received immunosuppressive therapy for at least three months prior to recruitment and had an eGFR ≥10 mL/min/1.73 $m^2$ | Recruited prior to biopsy | NA | NA | NA | Baseline: 0 Follow-up: 74.9 | Baseline: 0 follow-up: 12.8 | NA | Baseline: 36.6 Follow-up: 100 | NA |
| Barbour SJ. Kidney Int 2013 | Patients with biopsy-proven IgAN aged ≥18 years, with available follow-up data, recorded self-reported race, and non-missing proteinuria, eGFR, and mean arterial | The time of biopsy | 64 | 83 | NA | 18.3 | 21.3 | NA | 53.7 | NA |

| | | | | | | | | | | |
|---|---|---|---|---|---|---|---|---|---|---|
| | pressure at biopsy, excluding those with kidney failure at registration | | | | | | | | | |
| Coppo R. Kidney Int 2014 | Patients with biopsy-proven primary IgAN, ≥8 glomeruli in the renal biopsy specimen, and either a follow-up duration >1 year or progression to kidney failure within 1 year | Within 3 months of biopsy | 338 | 315 | NA | NA | NA | 10 | 39 | NA |
| Faucon AL. Nephrol Dial Transplant 2025 | Patients aged ≥18 years with non-dialysis CKD due to primary IgAN, registered in the Swedish Renal Registry between January 1, 2005, and December 31, 2021, and with available UACR measurements | Cohort enrollment | 399 | 133 | NA | 19.1 | 4.2 | NA | 83.1 | NA |
| Hirano K. Clin Exp Nephrol 2013 | Patients with IgAN who received 6 months of steroid therapy and had at least 1 year of follow-up | The time of initiation of steroid therapy | 80 | 22 | 23 | 100 | NA | NA | 44.0 | NA |
| Le W. Nephrol Dial Transplant 2012 | Patients with biopsy-proven primary IgAN aged ≥18 years at biopsy and with ≥12 months of follow-up | The time of biopsy | 527 | 353 | NA | NA | NA | NA | NA | NA |
| Lee K. Eur J Clin | Patients with biopsy-proven primary IgAN, eGFR ≥ 30 mL/min/1.73 m$^2$, proteinuria ≥ | Baseline was defined as 3 months | NA | NA | NA | 0 | 0 | 0 | 95.3 | NA |

| | | | | | | | | | | |
|---|---|---|---|---|---|---|---|---|---|---|
| Invest 2018 | 300 mg/day at the time of biopsy, at least 12 months of follow-up, and no history of immunosuppressive therapy | after starting RAS inhibitors for patients who initiated treatment post-biopsy; otherwise, it was the time of biopsy. | | | | | | | | |
| Liu J. Oncotarget 2017 | Patients with biopsy-proven primary IgAN, aged ≥18 years with an eGFR ≥ 15 mL/min/1.73 $m^2$, ≥ 36 months of follow-up, and at least eight glomeruli in the biopsy | The time of biopsy | 128 | 108 | NA | NA | NA | 43.8 | 84.5 | NA |
| Mohd R. PLoS One 2021 | Patients with biopsy-proven primary IgAN with minimum 1 year follow up | The time of biopsy | 36 | 42 | NA | NA | NA | 8.5 | 23.1 | NA |
| Nam KH. PLoS One 2014 | Patients with biopsy-proven primary IgAN, aged ≥18 years, with at least 12 months of follow-up and biopsy samples containing >7 glomeruli | The time of biopsy | 130 | 226 | NA | 11.0 | NA | NA | 78.0 | NA |
| Pitcher D. Clin J Am Soc | Patients aged ≥18 years with biopsy-proven primary IgAN, proteinuria >0.5 g/day or eGFR <60 mL/min/1.73 $m^2$, and at least 6 months of follow-up | The time of diagnosis | 215 | 175 | NA | NA | NA | NA | NA | NA |

| | | | | | | | | | | |
|---|---|---|---|---|---|---|---|---|---|---|
| Nephrol 2023 | | | | | | | | | | |
| Sarcina C. PLoS One 2016 | Patients with biopsy-proven IgAN enrolled in three randomized clinical trials | RCT enrollment | Baseline: 36<br>TAP: 58 | Baseline: 125<br>TAP: 144 | NA | 86.8 | 34.2 | NA | 50.5 | NA |
| Sato H. PLoS One 2021 | Patients with primary IgAN aged >20 years, newly diagnosed by renal biopsy, who were followed up for at least 5 years without receiving corticosteroids, other immunosuppressants, or tonsillectomy during the follow-up period | The time of biopsy | 43 | 10 | NA | 0 | 0 | 0 | 82.3 | NA |
| Shen X. Nephrol Dial Transplant 2025 | Patients with biopsy-proven primary IgAN aged ≥18 years, a follow-up duration of at least 1 year, not diagnosed with kidney failure at the time of biopsy, and with available follow-up data on 24-hour urinary protein and endpoint events | The time of biopsy | 293 | 680 | 296 | Follow-up: 43 | Follow-up: 26 | NA | Follow-up: 91 | NA |
| Sim JJ. Nephrol Dial Transplant 2025 | Patients aged ≥18 years with biopsy-proven primary IgAN, with at least 6 months of continuous the Kaiser Permanente Southern California membership prior to biopsy (allowing <45-day gaps) | The time of biopsy | 84 | 82 | NA | NA | NA | 40.8 | 69.8 | 0.3 |

| | | | | | | | | | | |
|---|---|---|---|---|---|---|---|---|---|---|
| Stamellou E. Clin Kidney J 2024 | Patients with biopsy-proven IgAN of European ancestry, aged 18–74 years, under nephrological care, and with either an eGFR of 30–60 ml/min/1.73 $m^2$ (corresponding to CKD stages G3, A1–3) or an eGFR ≥60 ml/min/1.73 $m^2$ with albuminuria (UACR >300 mg/g creatinine; corresponding to CKD stages G1–2, A3) | First study visit | 110 | 158 | NA | 15.1 | 5.9 | NA | 92 | NA |
| Takada D. Clin Exp Nephrol 2019 | Patients with biopsy-proven IgAN aged ≥16 years, with eGFR ≥30 mL/min/1.73 $m^2$, treated with tonsillectomy and steroid pulse therapy (<4 times), who had no history of renal transplantation and were followed for at least one | The time of biopsy | 72 | 52 | NA | 100 | NA | NA | NA | NA |
| Tan M. Kidney Blood Press Res 2015 | Adult patients with biopsy-proven primary IgAN who had an eGFR level >60 mL/min per 1.73 $m^2$, normal blood pressure, and a proteinuria level <1.0 g/24-h, >12 months of follow-up | The time of biopsy | 232 | 144 | NA | NA | NA | NA | Baseline: 61.4 Follow up: 77.1 | NA |
| Tanaka S. Clin J Am Soc | Patients with biopsy-proven primary IgAN, who had ≥10 | The time of biopsy | 266 | 163 | NA | NA | NA | NA | NA | NA |

| | | | | | | | | | | |
|---|---|---|---|---|---|---|---|---|---|---|
| Nephrol 2013 | glomeruli in biopsy specimens and available clinical data | | | | | | | | | |
| Tang C. Am J Kidney Dis 2024 | Patients with biopsy-proven primary IgAN, at least 2 visits, ≥12 months of follow-up, CKD stage <5 at first visit, and complete baseline data | The time of presentation | 127 | 351 | 139 | 40.1 | 33.0 | NA | 96.1 | NA |
| Xu R. Int J Nephrol Renovasc Dis 2025 | Patients with biopsy-proven primary IgAN recorded in the IgAN Database of Shenzhen Second People's Hospital, who had follow-up data for both urine protein-creatinine ratio and eGFR, a baseline eGFR ≥15 mL/min/1.73 $m^2$, and a follow-up duration of at least 6 months. | The time of biopsy | 225 | 114 | 88 | NA | NA | <0.3: 47.1 0.3-0.5: 47.7 0.5-1: 50.0 >1: 44.3 | <0.3: 78.2 0.3-0.5: 71.6 0.5-1: 76.3 >1: 67.2 | NA |
| Yu G. Front Med (Lausanne) 2022 | Patients with biopsy-proven IgAN with ≥6 months of follow-up and complete clinical data | The time of biopsy | 198 | 207 | NA | NA | NA | NA | NA | NA |

Abbreviations: CKD, chronic kidney disease; eGFR, estimated glomerular filtration rate; GC, glucocorticoid; IgAN, IgA nephropathy; IS, immunosuppressant; NA, not available; RAS, renin-angiotensin-aldosterone system; TAP, time-averaged proteinuria, RCT, randomized control trial; SGLT2, sodium glucose co-transporter 2; UACR, urinary albumin-to-creatinine ratio.

**Supplementary Table S2. Detailed scores of Newcastle-Ottawa Scale (NOS).**

| Study Name | Selection 1 | Selection 2 | Selection 3 | Selection 4 | Comparability a) | Comparability b) | Outcome 1 | Outcome 2 | Outcome 3 | NOS Stars | NOS Quality |
|---|---|---|---|---|---|---|---|---|---|---|---|
| Ai Z. Clin Nephrol 2020 | 1 | 1 | 1 | 1 | 1 | 1 | 1 | 1 | 0 | 8 | Good |
| Alexander S. Kidney Int Rep 2021 | 1 | 1 | 1 | 1 | 0 | 0 | 1 | 1 | 1 | 7 | Good |
| Barbour SJ. Kidney Int 2013 | 1 | 1 | 1 | 1 | 0 | 0 | 1 | 1 | 0 | 6 | Fair |
| Coppo R. Kidney Int 2014 | 1 | 1 | 1 | 1 | 0 | 0 | 1 | 1 | 0 | 6 | Fair |
| Faucon AL. Nephrol Dial Transplant 2025 | 1 | 1 | 1 | 1 | 1 | 1 | 1 | 1 | 1 | 9 | Good |
| Hirano K. Clin Exp Nephrol 2013 | 1 | 1 | 1 | 1 | 1 | 1 | 1 | 1 | 0 | 8 | Good |
| Le W. Nephrol Dial Transplant 2012 | 1 | 1 | 1 | 1 | 1 | 0 | 1 | 1 | 0 | 7 | Good |
| Lee K. Eur J Clin Invest 2018 | 1 | 1 | 1 | 1 | 1 | 1 | 1 | 1 | 0 | 8 | Good |
| Liu J. Oncotarget 2017 | 1 | 1 | 1 | 1 | 0 | 1 | 1 | 1 | 1 | 8 | Good |
| Mohd R. PLoS One 2021 | 1 | 1 | 1 | 1 | 0 | 0 | 1 | 1 | 0 | 6 | Fair |
| Nam KH. PLoS One 2014 | 1 | 1 | 1 | 1 | 1 | 1 | 1 | 1 | 0 | 8 | Good |
| Pitcher D. Clin J Am Soc Nephrol 2023 | 1 | 1 | 1 | 1 | 1 | 1 | 1 | 1 | 0 | 8 | Good |
| Sarcina C. PLoS One 2016 | 1 | 1 | 1 | 1 | 0 | 0 | 1 | 1 | 0 | 6 | Fair |

| | | | | | | | | | | |
|---|---|---|---|---|---|---|---|---|---|---|
| Sato H. PLoS One 2021 | 1 | 1 | 1 | 1 | 0 | 0 | 1 | 1 | 0 | 6 | Fair |
| Shen X. Nephrol Dial Transplant 2025 | 1 | 1 | 1 | 1 | 1 | 1 | 1 | 1 | 0 | 8 | Good |
| Sim JJ. Nephrol Dial Transplant 2025 | 1 | 1 | 1 | 1 | 1 | 1 | 1 | 1 | 0 | 8 | Good |
| Stamellou E. Clin Kidney J 2024 | 1 | 1 | 1 | 1 | 1 | 1 | 1 | 1 | 0 | 8 | Good |
| Takada D. Clin Exp Nephrol 2019 | 1 | 1 | 1 | 1 | 1 | 1 | 1 | 1 | 0 | 8 | Good |
| Tan M. Kidney Blood Press Res 2015 | 1 | 1 | 1 | 1 | 0 | 0 | 1 | 1 | 0 | 6 | Fair |
| Tanaka S. Clin J Am Soc Nephrol 2013 | 1 | 1 | 1 | 1 | 1 | 1 | 1 | 1 | 0 | 8 | Good |
| Tang C. Am J Kidney Dis 2024 | 1 | 1 | 1 | 1 | 1 | 1 | 1 | 1 | 0 | 8 | Good |
| Xu R. Int J Nephrol Renovasc Dis 2025 | 1 | 1 | 1 | 1 | 1 | 1 | 1 | 1 | 1 | 9 | Good |
| Yu G. Front Med (Lausanne) 2022 | 1 | 1 | 1 | 1 | 0 | 0 | 1 | 1 | 0 | 6 | Fair |

**Supplementary Table S3. Leave-one-out analyses for association between baseline proteinuria and adverse kidney outcomes.**

| Omitted study | HR (95%CI) | $I^2$, % |
|---|---|---|
| Ai Z. Clin Nephrol 2020 | 1.67 (1.31, 2.13) | 0.0 |
| Barbour SJ. Kidney Int 2013 | 1.71 (1.34, 2.19) | 0.0 |
| Faucon AL. Nephrol Dial Transplant 2025 | 2.00 (1.38, 2.89) | 0.0 |
| Hirano K. Clin Exp Nephrol 2013 | 1.73 (1.36, 2.20) | 0.0 |
| Sarcina C. PLoS One 2016 | 1.73 (1.36, 2.21) | 0.0 |
| Sim JJ. Nephrol Dial Transplant 2025 | 1.67 (1.30, 2.15) | 0.0 |
| Stamellou E. Clin Kidney J 2024 | 1.74 (1.31, 2.32) | 0.0 |
| Takada D. Clin Exp Nephrol 2019 | 1.88 (1.42, 2.48) | 0.0 |
| Tanaka S. Clin J Am Soc Nephrol 2013 | 1.71 (1.34, 2.17) | 0.0 |

Abbreviations: CI, confidence interval; HR, hazard ratio.

**Supplementary Table S4. Leave-one-out analyses for association between time-averaged proteinuria and adverse kidney outcomes.**

| Omitted study | HR (95%CI) | $I^2$, % |
|---|---|---|
| Le W. Nephrol Dial Transplant 2012 | 2.40 (1.25, 4.60) | 74.3 |
| Lee K. Eur J Clin Invest 2018 | 2.90 (1.43, 5.91) | 80.8 |
| Liu J. Oncotarget 2017 | 3.42 (1.61, 7.27) | 76.4 |
| Nam KH. PLoS One 2014 | 2.90 (1.42, 5.91) | 80.8 |
| Pitcher D. Clin J Am Soc Nephrol 2023 | 3.58 (1.84, 6.94) | 75.1 |
| Shen X. Nephrol Dial Transplant 2025 | 2.61 (1.22, 5.60) | 72.9 |
| Sim JJ. Nephrol Dial Transplant 2025 | 2.55 (1.23, 5.30) | 75.2 |

Abbreviations: CI, confidence interval; HR, hazard ratio.

**Supplementary Table S5. Leave-one-out analyses for association between time-averaged proteinuria and annual estimated glomerular filtration rate slope.**

| Omitted study | MD (95%CI) | $I^2$, % |
|---|---|---|
| Le W. Nephrol Dial Transplant 2012 | -0.80 (-1.39, -0.22) | 78.6 |
| Nam KH. PLoS One 2014 | -1.27 (-1.67, -0.87) | 49.2 |
| Pitcher D. Clin J Am Soc Nephrol 2023 | -1.01 (-1.71, -0.31) | 86.7 |
| Shen X. Nephrol Dial Transplant 2025 | -0.99 (-1.85, -0.13) | 86.3 |

Abbreviations: CI, confidence interval; MD, mean difference.
